# Feasibility of a livestream dance class for people with chronic stroke

**DOI:** 10.64898/2026.02.28.26347337

**Authors:** Sarah Gregman, Wade W Michaelchuk, Lauren C Belfiore, Kara K Patterson

## Abstract

**Background:** Adapted dance is a promising rehabilitation intervention for physical and psychosocial impairments in people with chronic stroke. However, in-person attendance is hindered by limited community ambulation, transportation, and schedule conflicts. At-home participation with a live-streamed dance program could address these issues, but psychosocial benefits may be diminished because of reduced social interactions. The primary objective of this study was to assess the feasibility and safety of a live-streamed dance program for chronic stroke. Secondary objectives were to characterize participants who choose live-stream vs in-person options and quantify pre-post changes in balance, gait and social connection.

**Method:** People with chronic stroke were given the choice of attending a live-streamed adapted dance program either in-person or at home twice a week for 4 weeks. *A priori* feasibility criteria were tracked, and participants were characterized with self-report (Center for Epidemiologic Studies Depression Scale; CES-D) and performance-based measures (e.g., Montreal Cognitive Assessment, Chedoke McMaster Assessment) at baseline. Pre-post measures of secondary outcomes included gait speed, Mini Balance Evaluation Systems Test (Mini-BESTest), Activities of Balance Confidence Scale (ABC), and Inclusion of Community in Self scale (ICS). Unpaired median/mean differences in baseline clinical presentation were used to compare in-person and live-stream participants. Paired median/mean differences were used to examine change in secondary outcomes with dance.

**Results:** Interest and enrollment rates for both groups combined were 87% and 38% respectively. Of the 13 people who enrolled, 8 chose in-person and 5 chose live-stream. In-person and live-stream attendance rates were 83% and 89% respectively, and retention rates were 80% and 75% respectively. At baseline, the in-person group had greater depressive symptoms (CES-D score, median [IQR] difference: 11.5 [-21.5, -5]), and faster mean gait speed (-25.8cm/s [-50.98, 0.006]) than the live-stream group. There were no pre-post changes in secondary outcome measures.

**Conclusions:** A live-streamed dance intervention featuring in-class and at-home participation is safe and feasible for people with chronic stroke. These results will inform a future randomized controlled trial to investigate the effects of a live-stream dance program with a longer duration while considering how factors such as gait function and mood may relate to the choice between in-person and at-home attendance.

## BACKGROUND

Stroke is a brain injury caused by the interruption of blood flow to the brain or bleeding around or into the brain due to a ruptured artery, and it is the leading cause of death and adult disability.(1) Stroke impacts various aspects of a person’s life including their mobility, speech, and cognition,(2) as well as their ability to participate in activities of daily living and reintegrate into their community.(3) Common lower extremity physical impairments include slow and asymmetrical gait, impaired balance, and increased risk for falls.(4, 5) Gains are made during inpatient rehabilitation, but people are left with persistent deficits at discharge, leading them to seek out alternative and long-term rehabilitation options.(4) Stroke also affects psychosocial well-being. After discharge home, people with stroke report symptoms of depression, social isolation,(6, 7) and moderate levels of stress.(8) Furthermore, approximately half of community-dwelling people with stroke have low balance confidence(9) which can lead to activity restriction, deconditioning and, consequently, worse quality of life and functional decline.(10, 11) Thus, stroke continues to affect the whole person even in the chronic stage and therefore, interventions that can address these persistent impairments are needed.

Dance is a worldwide human activity that involves the coordination of intentional movements synchronized to rhythmical stimuli, typically (though not exclusively) performed with other people.(12) Older adults who dance have more stable gait, faster motor reaction times, and better balance compared to older adults who do not dance.(13, 14) Some forms of dance have similar aerobic benefits to walking programs, and dance in general is associated with emotional and social well-being.(12, 15) Given the range of benefits, dance holds promise as a holistic intervention that can address both physical and psychosocial concerns expressed by people with chronic stroke. As a rehabilitation intervention, in-person dance programs for people with stroke in both the subacute and chronic stages are feasible and safe.(16-19) Several studies on dance with various neurorehabilitation patient populations report high levels of interest, retention, and adherence(16, 20) with some reporting that participants are eager to continue dancing upon completion of such dance programs.(16, 17) Preliminary evidence suggests dance improves gait and balance post-stroke.(16, 21) Finally, people with chronic stroke who completed a 10-week dance program reported that it moved beyond the physical benefits of typical exercise and conventional rehabilitation.(22) Specifically, the program provided a humanistic space that afforded opportunities to develop movement confidence, explore self-expression, and create social connections, thus confirming the psychosocial benefits of dance.(22) Creating social connections through dance is of interest given a recommendation from a 12-month study of post-stroke isolation that stated interventions should specifically foster connectedness to others and not just social support alone to address isolation.(23)

Most of the previous work on dance as a rehabilitation intervention has focused on in-person delivery.(20, 21) However, attending programs in person is not always practical or sustainable. Transportation and scheduling conflicts are common barriers to enrollment and adherence in a dance program by people with stroke.(16) Community ambulation (i.e., the ability to ambulate outside of the home) is achieved by approximately 60% of people with stroke.(24) This loss of community ambulation is associated with decreased participation in leisure activities and a loss of social contact.(6) Thus, stroke-related impairments prevent people from accessing community-based programs that may help improve those same impairments. Home exercise programs are one option to improve physical performance and address the barriers transportation and impaired community ambulation may present. However, exercises performed by an individual with either instructional videos or a paper-based exercise prescription lack the social interactions afforded by in-person dance-based interventions and thus may not provide the same psychosocial benefits. A potential solution is a live-streamed dance program that allows for synchronous participation at home. Previous qualitative work found that people with stroke, physiotherapists, and dance teachers are in favour of live-streamed dance classes to address accessibility barriers, and promote long-term engagement in dance.(25)

The primary objective of this study was to assess the feasibility and safety of a live-streamed dance program for people with chronic stroke. The secondary objectives were to 1) characterize participants who select in-class and at-home delivery of the dance program and 2) compare changes in social connectedness, balance, and gait with the program when delivered at home compared to in-person. Information about feasibility, safety, interest, and potential effects can inform the design of a future randomized controlled trial of live-streamed dance programs for people with stroke.

## METHODS AND MATERIALS

### Participants

People with stroke living in the community were eligible to participate if they were: a) >6 months post stroke, b) able to walk ≥10 meters without assistive devices, c) cleared to exercise by their physician, and d) able to tolerate ≥50 minutes of physical activity with rests as needed. Participants were excluded from participation if they had moderate or severe self-reported hearing loss or other diagnosed conditions that affected balance and gait (e.g., osteoarthritis or Parkinson’s disease). Participants were recruited from the surrounding community through printed poster advertisements in a rehabilitation hospital and private physiotherapy clinics. Digital advertisements were also posted through social media and websites of non-profit stroke-related organizations. Finally, potential participants in a research volunteer database were also contacted. The Research Ethics Board at the University Health Network approved this study, and all participants provided written informed consent. The research was conducted in accordance with the TriCouncil Policy Statement: Ethical Conduct for Research Involving Humans.(26)

We aimed to recruit a maximum of 10 people for the in-person group and 5 people for the live-stream group. This sample size was determined by resource constraints as well as practical and safety considerations.(27) The funding amount and timeline allowed for the adapted dance program to be delivered once which limited the sample size to 1 class cohort. The size of the class cohort was determined by practical and safety considerations: the maximum number of participants that would allow for 1-on-1 attention and feedback as well as physical support and/or standby assistance from 1 teacher and 2-3 dance assistants.

#### Dance class structure and content

The live-streamed dance intervention was informed by previous work on structuring post-stroke dance programs(25) and a feasibility study of a 10-week in-person post-stroke dance program.(16) The program duration was reduced to 4 weeks to address the primary objective to examine the feasibility of live-stream delivery while recognizing it was likely insufficient to induce changes in physical performance. A dance instructor with experience teaching people with disabilities delivered classes in person, twice a week. Classes were 50 minutes in length and included a seated warm-up, chair/standing choreography, a 10-minute break, dance moving across the floor, centre floor choreography or partnered dancing, and a closing bow. The program incorporated individual and group dance activities, and a variety of styles (e.g., salsa, ballet, waltz, partnered, and non-partnered) and music genres (e.g., American standards, Latin, etc.) to meet different participants’ interests.(25) More detail about the dance class content and structure can be found in Patterson and colleagues.(16) Two or 3 dance assistants were available at each class to support participants in the classroom and at home.

#### Classroom set-up and live-stream delivery

At the time of enrollment, participants were given the option to attend classes in person or remotely via live stream. All participants attended the first class in person and the remaining 7 of the 8 in-person classes were live-streamed using a wide-angle webcam (MeetUp™, Logitech International, Lausanne, Switzerland) and Zoom (Zoom Video Communications Inc., San Jose, California, USA). The in-person dance classroom was set up with chairs in a circle with the camera positioned within that circle as an additional “member” of the class to create a sense of presence for the at-home group. Figure 1 includes images from the in-person and live stream setup. The dance instructor intentionally spoke to the camera during each dance activity to ensure the at-home group felt “seen”. One dance assistant was always stationed in front of the camera to demonstrate the dance exercises and activities directly to the at-home participants.

**Figure 1.**
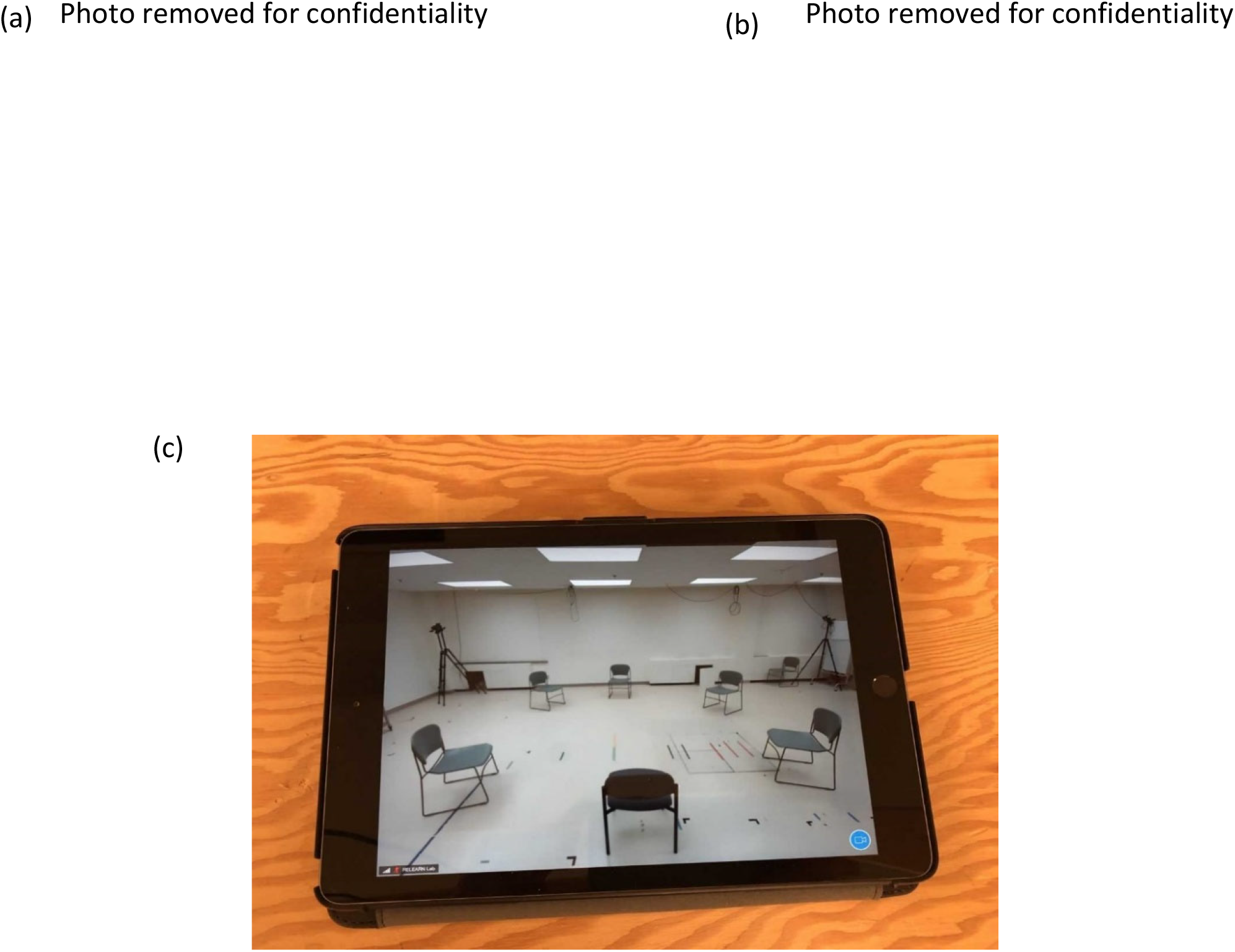
Dance program set up. a) In-person participants performing ballet plié warm-up exercise and a dance assistant demonstrating the exercise to the camera for the live-stream participants, b) in-person participants performing partnered dancing with the dance assistant demonstrating simulated partner dancing with a scarf and chair for the live-stream participants, c) tablet display used by live-stream participants.

#### Equipment, training, safety, and technical support for at-home participation by live-stream

Participants in the live-stream group attended the first class in person to foster connections with the dance instructor, dance assistants, and other study participants. This also allowed for confirmation of their ability to perform class movements and exercises safely. After the first class, the live-stream participants received tablets (iPad, Apple, Cupertino, California, USA) with the Zoom application pre-installed. A research team member provided live-stream participants with 1-on-1 training on using the tablet, including opening Zoom and connecting to the live-stream class.

After the first in-person class and before the remaining 7 live-streamed classes, these participants underwent a 15-30 minute virtual review of their home space with a registered physiotherapist. Using the tablet provided, live-stream participants met with the physiotherapist to give them a virtual tour of the space in their home where they would dance. The physiotherapist asked the participant to provide an estimation of the available floor area, to view the flooring and any furniture in the space such that hazards could be identified. The PT provided suggestions such as removing loose mats or rugs and re-organization of furniture to ensure sufficient space, as well as education about safe participation in the dance class at home.

During the live-streamed dance classes, a research team member was present to address technical difficulties. Throughout each class, the research team member monitored all participants on the Zoom platform to answer questions, troubleshoot technical difficulties, and respond to any potential emergency situations.

### Data Collection Protocol

#### Feasibility and Safety

The present study was guided by work on feasibility studies by Bowen and colleagues(28) Who outlined the purpose of feasibility studies as: “to identify not only what—if anything—in the research methods or protocols needs modification but also how changes might occur”(28). Bowen and colleagues describe 8 areas of focus addressed by feasibility studies and 4 of them applied to the current work: acceptability, demand, implementation and limited efficacy testing Eight domains within these 4 areas of focus were identified *a priori* as important for intervention implementation and a subsequent randomized controlled trial.

1. *Interest*: the number of participants interested in the study expressed as a percentage of the total number of participants approached
2. *Enrolment*: the number of participants enrolled in the study expressed as a percentage of all participants screened for the study. Reasons for not enrolling were also tracked.
3. *Interest in live-stream option*: the number of enrolled participants who chose the live-stream option, expressed as the percentage of all participants enrolled in the study
4. *Retention*: the number of people who completed the study expressed as a percentage of all participants enrolled in the study
5. *Attendance*: the number of classes attended expressed as a percentage of the number of classes the participant was expected to attend
6. *Adverse events*: the number of major adverse events defined as falls causing serious injury (e.g., fractures); events requiring medical attention; or side effects other than those expected with moderate exercise (e.g., fatigue, delayed onset muscle soreness), and the number of minor events that affected attendance or participation (e.g., fatigue)
7. *Technical issues*: the number of events requiring assistance from the research team member or that prevented attendance
8. *Participant satisfaction*: assessed with a 16-item exit survey that asked participants to rate 10 statements about the dance program using a 5 point scale (1 = ‘strongly disagree’ and 5 = ‘strongly agree’) and 6 open-ended questions: what they liked best and least about the program, recommendations for improving the dance program, would they would choose the other class option (i.e., in-class or live-stream), and how much they would pay per class if the program was not free. This survey was administered by a research physiotherapist at the in-person post-intervention assessment within 1-8 days after the final class.

#### Secondary Outcome Measures

All participants attended an in-person assessment within 1-12 days before/after the dance intervention to collect clinical measures and secondary variables of interest. The measures were administered by 2 research physiotherapists who were not involved in the dance classes and were blinded to the participants’ group.

a. *Clinical Characteristics:* Demographics (age, sex, stroke onset) were collected with a questionnaire. Stroke severity was measured with the National Institutes of Health Stroke Scale (NIHSS)^13^ and foot and leg motor impairment were measured with the Chedoke McMaster Stroke Assessment (CMSA).(29) Cognitive impairment was measured with the Montreal Cognitive Assessment (MoCA),(30) and depression was measured with the Center for Epidemiological Studies Depression (CES-D).(31)
b. *Secondary variables of interest:* The Inclusion of Community in Self (ICS) scale was used to measure social connection with the group. The ICS scale is a set of 5 figures featuring 2 circles of increasing overlap. One circle represents the self, and the other represents the dance class. Participants are asked to select the figure that represents their connection to the dance class. Greater feelings of connectedness are represented by circles with greater overlap.(32) The ICS was selected because it differentially assesses connectedness to the community (in this case the dance class), distinct from close relationship connectedness.(32) Balance was evaluated using the Mini-Balance Evaluation Systems Test (Mini-BESTest),(33, 34) and balance confidence was measured with the Activities of Balance Confidence (ABC) scale.(35, 36) Spatiotemporal gait parameters were measured using a pressure-sensitive mat (Protokinetics, Havertown, PA, USA).(37) At least 18 footfall events were collected during preferred pace walking to ensure reliability.(37) For the purposes of this feasibility study, the analysis focused on gait speed as a measure of overall gait function.(38)

#### Qualitative and Quantitative Data Processing and Statistical Analysis

Gait data collected with the pressure sensitive mat was processed by a research team member blinded to group assignment. Descriptive statistics were used to summarize participant demographics and clinical characteristics. To address the primary objective, data for the 8 feasibility domains were summarized and reported as frequency counts and percentages for quantitative variables, and descriptive summaries with frequency counts for qualitative variables. Descriptive content analysis was used to characterize participant responses to the open-ended questions on the satisfaction questionnaire. Common ideas and concepts were identified by one research team member and verified independently by a second team member. The secondary objectives were addressed with estimation statistics, which are a group of methods that focus on the estimation of effect sizes and their confidence intervals.(39) To address objective 2a, between-group comparisons at baseline were used to characterize people who choose in-class and live-stream options using unpaired median and mean differences, and data were plotted as Gardner-Altman plots.(40) All participants were included in this baseline analysis, including the participants who eventually withdrew. Objective 2b was addressed by examining within-group differences from pre-to post-dance program for secondary variables of interest (ICS, Mini-BESTest, ABC, and gait speed) using paired median and mean differences. Data were also plotted as Cumming estimation plots.(40) Only data from participants who completed the program were included in this analysis. P-values were calculated for legacy purposes.

## RESULTS

### Participants

Of the 34 people screened, 11 people did not meet the eligibility criteria, 10 people did not enroll (see Table 2 for reasons), and 13 people did enroll in the program. Eight participants chose the in-person option and 5 chose the live-stream option. All participants in the live-stream option were assessed as physically safe to participate from home and their home space was deemed suitable for dancing by the physiotherapist. Ten participants completed the program and 1 participant in the in-person and 2 participants in the live-stream group withdrew themselves from the study. The baseline demographics and clinical presentation of the 13 enrolled participants are summarized in Table 1.

**Table 1.**
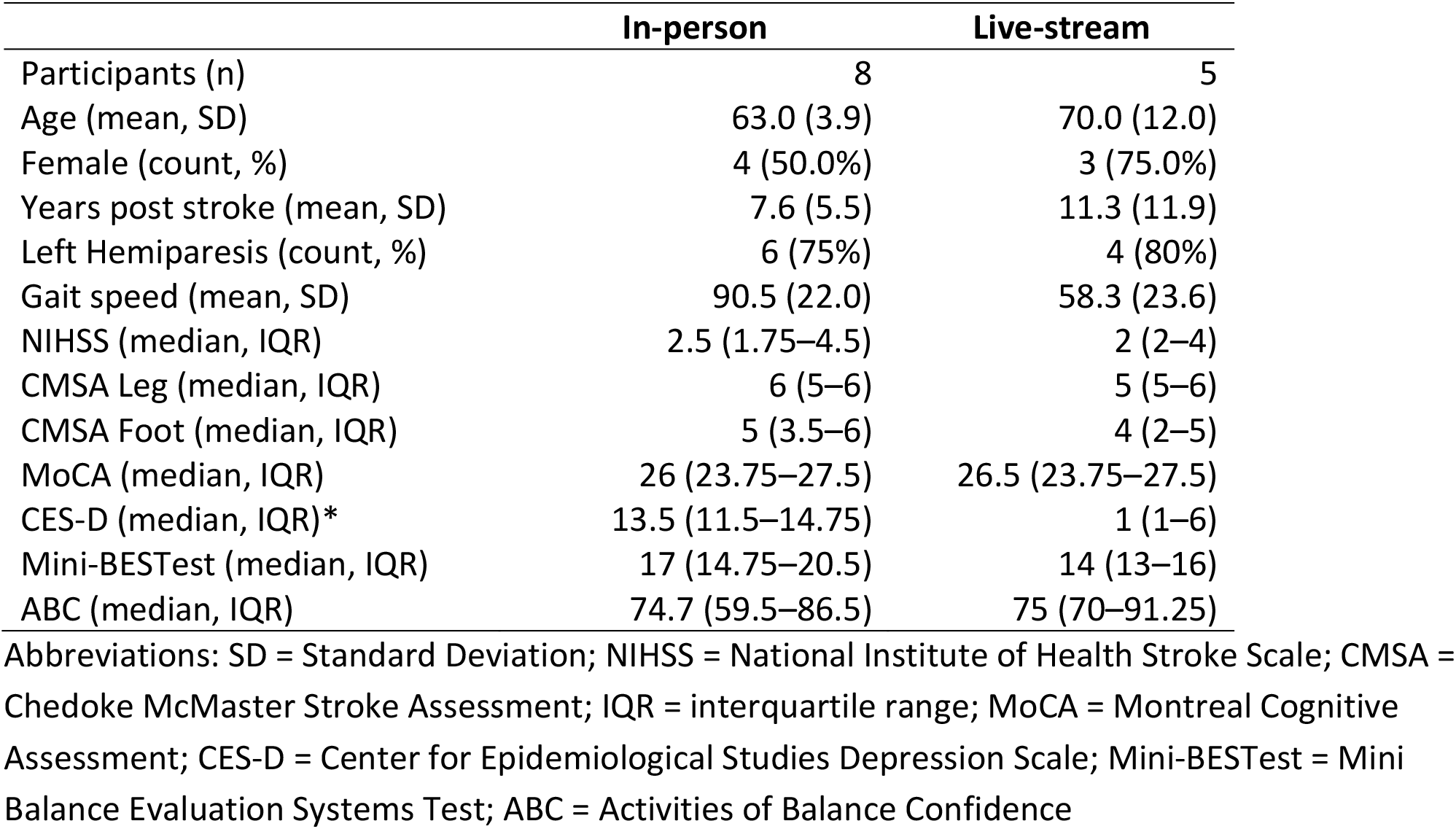
Baseline participant demographics and clinical presentation.

|  | In-person | Live-stream |
| --- | --- | --- |
| Participants (n) | 8 | 5 |
| Age (mean, SD) | 63.0 (3.9) | 70.0 (12.0) |
| Female (count, %) | 4 (50.0%) | 3 (75.0%) |
| Years post stroke (mean, SD) | 7.6 (5.5) | 11.3 (11.9) |
| Left Hemiparesis (count, %) | 6 (75%) | 4 (80%) |
| Gait speed (mean, SD) | 90.5 (22.0) | 58.3 (23.6) |
| NIHSS (median, IQR) | 2.5 (1.75–4.5) | 2 (2–4) |
| CMSA Leg (median, IQR) | 6 (5–6) | 5 (5–6) |
| CMSA Foot (median, IQR) | 5 (3.5–6) | 4 (2–5) |
| MoCA (median, IQR) | 26 (23.75–27.5) | 26.5 (23.75–27.5) |
| CES-D (median, IQR)* | 13.5 (11.5–14.75) | 1 (1–6) |
| Mini-BESTest (median, IQR) | 17 (14.75–20.5) | 14 (13–16) |
| ABC (median, IQR) | 74.7 (59.5–86.5) | 75 (70–91.25) |

Abbreviations: SD = Standard Deviation; NIHSS = National Institute of Health Stroke Scale; CMSA = Chedoke McMaster Stroke Assessment; IQR = interquartile range; MoCA = Montreal Cognitive Assessment; CES-D = Center for Epidemiological Studies Depression Scale; Mini-BESTest = Mini Balance Evaluation Systems Test; ABC = Activities of Balance Confidence

**Table 2.** Feasibility criteria.

| Feasibility domain | Description | Result |
| --- | --- | --- |
| 1) Interest in dance | number of participants interested in the study expressed as a percentage of the total number of participants approached about the study | 87% (34/39) |
| 2) Enrolment rate | number of participants enrolled in the study expressed as a percentage of all participants screened for the study. Reasons for not enrolling were also tracked. | 38% (13/34)<br>[Reasons for not participating scheduling issues (n=6); technology concerns (n=1); transportation and stamina concerns (n=1); previous injury (n=1); lack of interest (n=1); ineligible (n=11)] |
| 3) Interest in live-stream option | number of enrolled participants in each option, expressed as the percentage of all participants enrolled in the study | In-person 62% (8/13)<br>Live-stream 38% (5/13) |
| 4) Retention | number of people who completed the study expressed as a percentage of all participants enrolled in the study | 77% (10/13)<br>In-person: 75% (6/8)<br>Live-stream: 80% (4/5) |
| 5) Attendance | number of classes attended expressed as a percentage of the number of classes the participant was expected to attend | In-person: 83 (18)%<br>Live-stream: 89 (11)%<br>[Reasons for absence: scheduling conflicts, illnesses, poor weather, and transportation issues (in-class only)] |
| 6) Adverse events | number of adverse events defined as falls causing serious injury (e.g., fractures); events requiring medical attention; or side effects other than those expected with moderate exercise (e.g., fatigue, delayed onset muscle soreness) | Major: 0<br>Minor: 4<br>[Minor events included: fatigue unrelated to the intervention that prevented attendance (n=3); muscle soreness as expected with exercise (n=1)] |
| 7) Technical issues | number of events requiring assistance from the research team member or that prevented attendance | 15<br>[Issues included: difficulty with initial sign-on to Zoom and audio/video connectivity issues, internet connectivity, low battery.] |

\*Note: Two at-home participants could only attend classes once per week—an arrangement pre-approved by the study team prior to initiating the intervention. Attendance was tracked with respect to the classes participants were expected to attend, and therefore, the expected attendance for these 2 participants was 4 classes.

### Objective 1: Feasibility and safety of a synchronous live-stream dance program

The results for 7 of the 8 feasibility and safety domains are summarized in Table 2. There were no major adverse events observed during the classes, but 1 person reported a fall outside of the dance class and chose to participate in the classes in a seated position for 1 week.

The results for domain 8, participant satisfaction, are summarized in Figure 2. All participants who withdrew agreed to complete the exit questionnaire, and the live-stream participant who withdrew also agreed to complete the post-program outcome assessment. There was at least one technical issue that occurred during each class. All technical issues were dealt with by the research team member who was assigned to monitor the live-stream, and according to informal observations they were resolved without any disruption to the class flow or the other participants. For the questionnaire item “overall I enjoyed the dance class”, the unpaired median difference between the in-class and live-stream responses was 0 [-1, 0] (p=0.72). For the item “if I could, I would continue participating” the unpaired median difference was -1 [-2, -1] (p=0.69).

**Figure 2.**
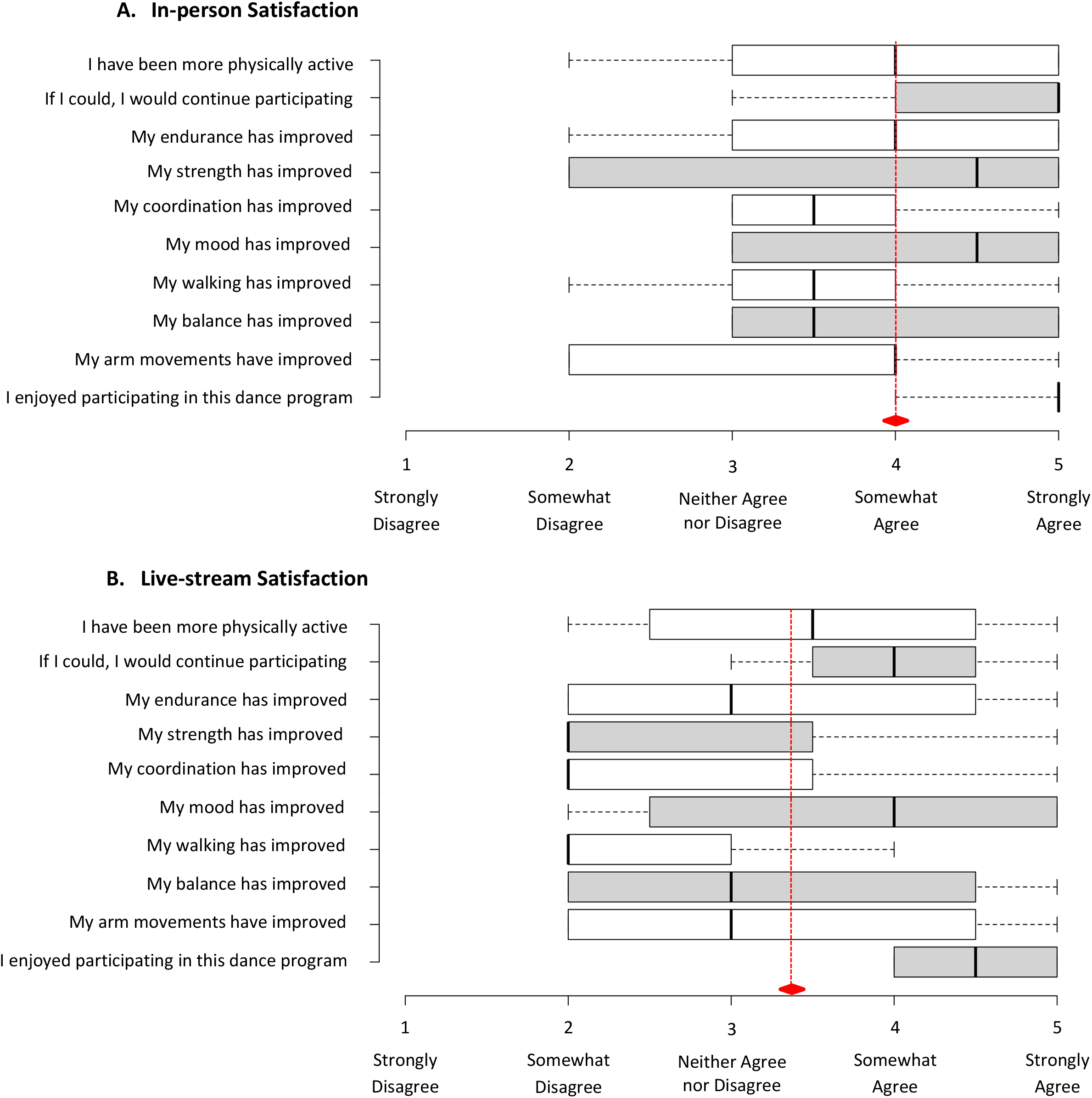
Exit questionnaire responses. Ratings of satisfaction with the dance program for the a) in-class and b) live stream groups. Solid black lines represent the median scores, edges of the box plots represent the interquartile range, and the whiskers represent minimum and maximum values reported. Alternating grey and white bars are used to facilitate distinguishing between items. Red diamonds indicate average overall satisfaction calculated as median of responses to all questions.

Through descriptive content analysis the following common concepts and ideas were identified. The aspect participants in both groups liked best about the program was the dancing itself. Socializing and physical improvements (e.g., balance and movement fluidity) were additional benefits identified by in-class participants. Aspects of the program that participants from both groups liked least included too much talking and sitting. Both groups suggested the dance program could be improved by increasing the time spent dancing and addressing some technical issues, such as larger screens for the live stream and a microphone for the instructor. Three of the 8 in-person participants (38%) reported interest in the live-stream option because it addresses issues with travel and it appeared more convenient. The 5 in-person participants not interested in the live-stream option cited motivation and socialization associated with the in-person option, and technical issues with the live-stream as reasons to avoid this option. Two of the 4 live-stream participants (50%) reported they would like to try the in-person option because it looked like fun. The convenience of at-home participation and difficulty with travel were the reasons the other 2 live-stream participants (50%) indicated they would not try the in-person option. Seven in-person (88%) and 3 live-stream participants (75%) indicated they would pay for the dance program, with most willing to pay CAD 5/class and some willing to pay up to CAD 20/class.

### Objective 2a: Characterize people who choose in-person vs live-stream dance

The baseline values for demographic and clinical measures (Table 1) were similar between the in-person and live-stream groups except for CES-D scores and gait speed (Figure 3). The in-person group had higher CES-D scores compared to the live-stream group with a mean difference [95% CI] of -11.5 [-21.5, -5], p=0.02). The in-person group had faster gait speed at baseline with a mean difference [95% CI] of -25.8 [-50.98, 0.006], p=0.09.

**Figure 3.**
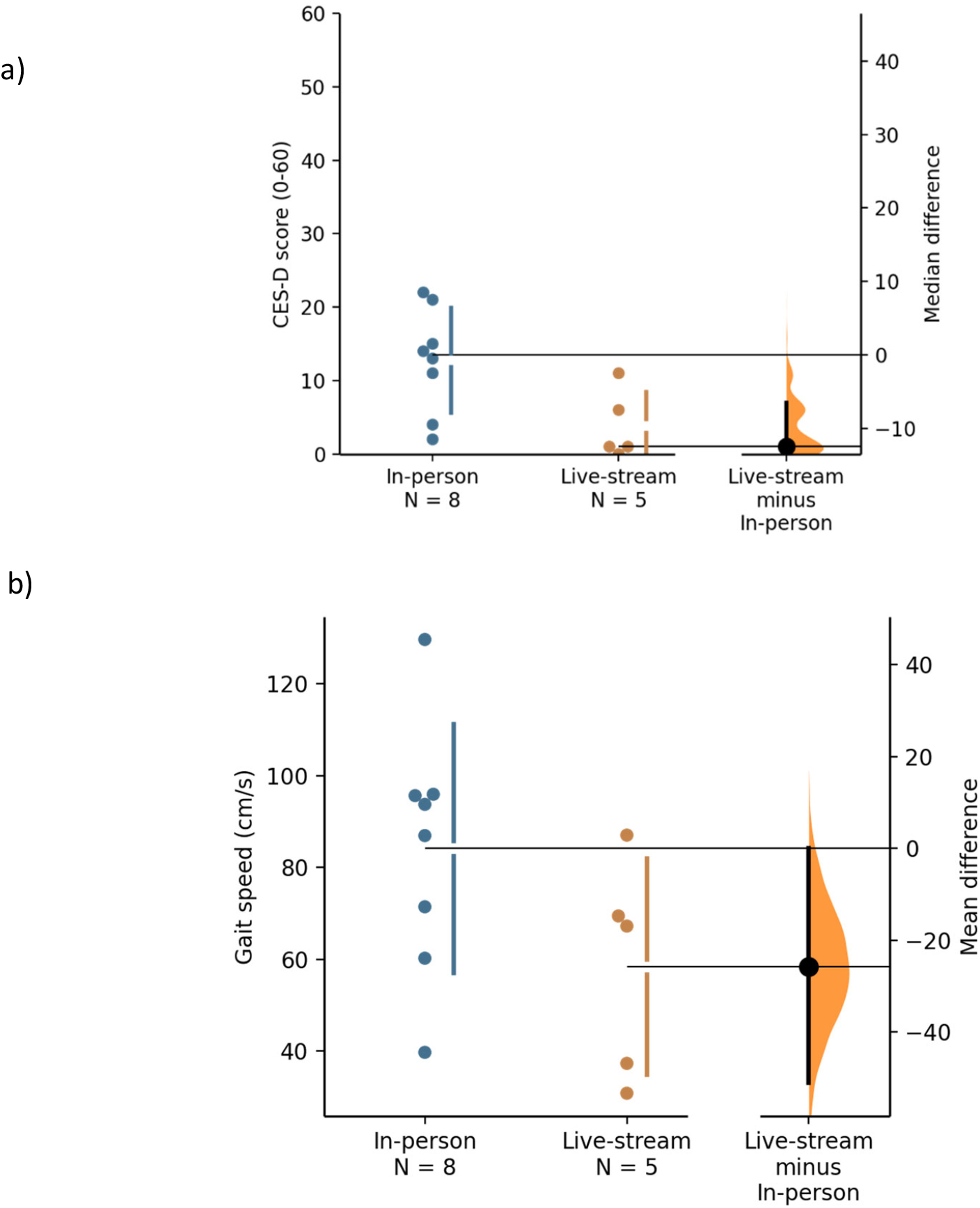
Baseline clinical presentation. The median difference in CES-D between the in-person and at-home groups illustrated in a Gardner-Altman estimation plot^35^ for (a) CES-D and (b) gait speed. Both groups are plotted on the left axes, the median difference is plotted on the right axis as a bootstrap sampling distribution. The median difference is a dot and the 95% CI is indicated by the ends of the vertical error bar.

### Objective 2b: Compare pre-post- dance class changes in secondary outcome measures between in-person and live-stream groups

Pre- and post-values of secondary outcome measures for the in-person and live-stream participants who completed the program are summarized in Table 3. None of the paired mean and median differences from pre-to post-dance program were significant for either group.

**Table 3.** Secondary Outcome Measures.

|  | Pre dance program |  | Post dance program |  |
| --- | --- | --- | --- | --- |
|  | In-person (n=6) | Live-stream (n=4) | In-person (n=6) | Live-stream (n=4) |
| ICS (0-5) | 3.5 [4–4.8] | 1.5 [1–2] | 4.5 [3.3–5] | 4.5 [3.3–5] |
| Mini-BESTest (0-28) | 17.5 [15–21.5] | 15 [13.8–17.5] | 18.5 [17.3–19.8] | 20 [14.5–20] |
| ABC Scale (0-100%) | 74.7 [63.6–86.5] | 83.1 [70.9–91.7] | 78.4 [62,4–91.9] | 85.6 [64.1–87.2] |
| Speed (cm/s) | 86.2 (29.7) | 65.2 (20.6) | 86.7 (27.1) | 61.1 (24.8) |

Values presented are mean and standard deviation (SD) or median and interquartile range [IQR]. ICS = Inclusion of Community in Self; Mini-BESTest = Mini-Balance Evaluation Systems Test; ABC Scale = Activity-specific Balance Confidence scale

## DISCUSSION

The main finding of this study is that a live-streamed post-stroke dance program featuring synchronous participation with people in class and at home was safe, feasible, and well-attended. Overall enjoyment of the program was similar between the 2 groups, but the in-person participants were more likely to want to continue with the program, indicating this may be the preferred format. Furthermore, baseline differences in clinical presentation highlight potential factors that could influence the choice of in-person vs live-stream dance.

Our previous work on an in-person only dance program for people with chronic stroke(16) is an appropriate comparator for the present hybrid dance program, particularly because the same dance instructor taught both programs and the structure and content of the classes were similar. This comparison provides insight into the advantages and challenges of offering the at-home live-streamed option. Interest in dance was similar between the current hybrid program and the previous in-person-only program (87% vs 90.9%, respectively), but the enrollment rate was lower (38% vs 66.7% respectively).(16) Similar to the previous in-person program, the most common reason for not enrolling in the present hybrid program was scheduling conflict. The attendance rate in the present study was similar for the in-class and live-stream groups (83% and 89%, respectively), but was lower compared to the previous in-person-only program (92.5%).(16) The lower attendance record for the present hybrid program is curious, given that the demand on the participants (4 weeks, 8 classes) was less than the previous in-person program (10 weeks, 20 classes). It is unclear whether the live-stream option altered the dance class experience in such a way as to be detrimental to attendance. However, it is worth noting that, despite being lower than previous in-person only dance program, compared to other in-person group exercise programs for people with chronic stroke (e.g., 75%(41) and 81%(42)), class attendance was strong.

In general, the in-person option appears preferable to the at-home live-stream option for dancing. A greater proportion of people (62%) chose the in-person over the live-stream option (38%) at baseline. Furthermore, at the end of the program, after experiencing one option and observing the other option, participants were asked if they would consider switching if the dance program were to continue. Only 38% of the participants who danced in person were interested in trying the live-stream option, while 50% of the participants in the live-stream option were interested in switching. Across both groups, common reasons for preferring in-person dancing were related to the social nature of dancing in a group. Common concerns that reduced the desire to switch to the live-stream option were technology-related. However, future dance programs may consider providing a live-stream option, despite the preference for in-person, for the sake of program convenience and accessibility. In the present study, participants appreciated the convenience and the reduced burden associated with traveling provided by the live-stream option.

Despite the small sample size, there were interesting baseline differences in mood and gait function between the in-person and live-stream groups that warrant discussion. These differences highlight factors that may impact the design and implementation of future live-stream dance programs and should be considered in future investigations. The in-person group had more depressive symptoms as measured by the CES-D, and 2 participants had scores >20, which is the cutoff for depression.(43) No participants in the live-stream group had a CES-D >20. This is relevant considering the relationship of depression and social isolation post-stroke. Social isolation (reported by ∼50% of people post-stroke(23)) is associated with an increase in depressive symptoms and limitations in activities of daily living.(44). Social and recreational interactions foster feelings of connectedness with others(23) and people post-stroke seek these opportunities to counteract isolation. (23) It is possible that some participants in the present study chose the in-person dance option for this reason. Social isolation was not measured in the present study; thus, firm conclusions cannot be drawn from this data. However, the group differences in depressive symptoms warrant further investigation. In line with the recommendation for individualized care,(23) it may be more appropriate to recommend in-person over live-stream dancing for a person post-stroke based on their level of social isolation and depressive symptoms.

Another difference at baseline was that the live-stream group had slower gait than the in-person group. This difference did not reach statistical significance; however, the magnitude of the mean difference and its 95% CI (-26cm/s, [-51; 0.1]), suggest that it is meaningful. Gait speed is an important indicator of the capacity for community ambulation.(45) At baseline, the mean speed exceeded the threshold for full community ambulation (0.8m/s (45)) for the in-person group but not the live-stream group. It is possible that participants with less capacity for community ambulation chose the live-stream option because it allowed them to participate in dance while avoiding the barriers that travel and moving in the community might pose.

Participants did not show significant improvement in secondary measures of balance, balance confidence, connection, or gait speed, which was expected given the duration of the program was shortened for the purposes of this feasibility study. The present results suggest it is safe and feasible to deliver a synchronous, live-streamed dance program for people with chronic stroke, and further investigation is warranted. It is important to emphasize the multiple measures taken to ensure the safety of all participants, which likely contributed to the absence of major adverse events. For the in-person group, dance assistants were available to provide standby assistance, which enabled participants to explore different movement ranges and patterns they may not have been safe to do independently. For the live-stream group, the virtual appointment with a physiotherapist ensured their home environment was safe to dance in. Furthermore, the researcher dedicated solely to monitoring the live-stream participants through Zoom also ensured that a quick response was possible if unsafe or emergency situations arose. It is strongly recommended that future work investigating live-stream dance includes these measures to achieve the 0 adverse event rate observed in the present study. Future studies should investigate the effects of such a program with a duration (∼10-12 weeks) which is more likely to produce improvements in physical and psychosocial domains.

## Limitations

This feasibility study focused on people with chronic stroke living in the community in a large Canadian city and thus the results may not be generalizable to people in the subacute stage, people in inpatient rehabilitation, or people in other living situations or rural areas. The short duration of the dance program was a practical decision to facilitate an investigation of feasibility and safety, but it limited the ability to detect change in physical and psychosocial outcomes. The small sample size is typical for feasibility studies which are intended to test an intervention in a limited way (i.e., convenience sample, intermediate outcomes, shorter follow-up period) and with limited statistical power.(28) Thus, while firm conclusions cannot be drawn from these preliminary results, they will inform a future randomized controlled trial and highlight additional avenues for investigation (i.e., baseline differences).

## Conclusions

In conclusion, a live-stream dance program for people with chronic stroke, living in the community is safe and feasible. In-person dance may be the preferred option, particularly for those with depressive symptoms. However, providing the option for attendance at home by live-streaming the class (with appropriate safety measures) may ensure inclusion and accessibility for those with limited capacity for community ambulation.

## Data Availability

The datasets generated and analyzed during the current study are not publicly available due to research ethics and privacy considerations

## LIST OF ABBREVIATIONS

ABC: Activities of Balance Confidence Scale
CES: Center for Epidemiological Studies-Depression Scale
CMSA: Chedoke McMaster Stroke Assessment
ICS: Inclusion of Community in Self scale
Mini-BESTest: Mini-Balance Evaluation Systems Test
MoCA: Montreal Cognitive Assessment
NIHSS: National Institutes of Health Stroke Scale

## ETHICS APPROVAL AND CONSENT TO PARTICIPATE

This study was approved by the University of Toronto Ethics Board (Protocol #34233) and the University Health Network Ethics Board (Protocol #16-6018) All participants provided informed consent and the research was conducted in accordance with the TriCouncil Policy Statement: Ethical Conduct for Research Involving Humans.

## COMPETING INTERESTS

None to declare

## FUNDING

This study was supported by funding from the Canadian Partnership for Stroke Recovery Catalyst Grant (no grant number available)

## AUTHORS CONTRIBUTIONS (CRediT taxonomy)

**Sara Gregman:** formal analysis, writing-original draft, writing-review & editing, visualization; **Wade W Michaelchuk:** formal analysis, investigation, writing-original draft, writing-review & editing, **Lauren Belfiore:** investigation, data curation, writing-review & editing, visualization, project administration, **Kara K Patterson:** conceptualization, methodology, formal analysis, investigation, resources, writing-original draft, writing-review & editing, visualization, supervision, funding acquisition

## Notes

### Competing Interest Statement

The authors have declared no competing interest.

### Author Declarations

Ethics committee of the University Health Network gave ethical approval for this work.

